# Awareness and Willingness to Use HIV Infection Pre-Exposure Prophylaxis among Rwandan Men Who Have Sex with Men: Findings from a Web-based Survey

**DOI:** 10.1101/2023.11.01.23297747

**Authors:** Athanase Munyaneza, Viraj V. Patel, Nataly Rios Gutierrez, Qiuhu Shi, Benjamin Muhoza, Gallican Kubwimana, Jonathan Ross, Etienne Nsereko, Gad Murenzi, Laetitia Nyirazinyoye, Leon Mutesa, Kathryn Anastos, Adebola Adedimeji

**Affiliations:** Einstein-Rwanda Research and, Capacity Building Program, Research for Development (RD Rwanda), Kigali, Rwanda; School of Public Health, College of Medicine and Health Sciences, University of Rwanda, Kigali, Rwanda; Division of General Internal Medicine, Montefiore Medical Center, Bronx, New York, USA; New York Medical College, Valhalla, New York, USA; Center for Human Genetics, College of Medicine and Health Sciences, University of Rwanda, Kigali, Rwanda; Department of Epidemiology and Population Health, Albert Einstein College of Medicine, Bronx, New York, USA

**Author notes:** Corresponding Author: Munyaneza Athanase.

**Keywords:** Men who have sex with men, Pre-exposure prophylaxis, Web-based survey, Rwanda, Sub Sahara Africa

## Abstract

**Introduction:** Pre-exposure Prophylaxis (PrEP) is a daily pill intended to reduce the risk of acquiring Human Immunodeficiency Virus (HIV) when taken as prescribed. It is strongly recommended for Men who have sex with Men (MSM) at high risk of HIV transmission to minimize infection risk. Despite its proven effectiveness, there is a lack of information about awareness and willingness to use PrEP among Rwandan MSM. In the context of HIV acquisition, the purpose of this study was to ascertain the awareness and willingness to use PrEP among high-risk Rwandan MSM. The findings of this research will provide valuable perspectives to mold policy and direct the effective execution of PrEP within the country.

**Method:** This is a cross-sectional study design that utilized a web-based survey conducted between April and June 2019 to assess awareness and willingness to use PrEP among sexually active MSM in Rwanda. A snowball sampling technique was used to recruit participants who were contacted via social medial such as WhatsApp and e-mail. To be eligible, participants were supposed to be sexually active, aged ≥18 years, self-identify as MSM, residence in Rwanda, self-reported engagement in receptive or insertive anal sex in the last 12 months, and self-reported HIV-negative sero-status. We assessed two primary outcomes: PrEP awareness (having ever heard of PrEP) and willingness to use PrEP within one month of completing the survey. Multivariable logistic regression was performed to identify participant characteristics associated with PrEP awareness and willingness to use it.

**Results:** Among the 521 participants included in the analysis, 63% were aged below 24 years. The majority (73%) demonstrated awareness of PrEP. Factors associated with PrEP awareness included residing outside of the capital, Kigali, as opposed to living in Kigali (adjusted odds ratio [aOR] 2.35, 95% confidence interval [CI] 1.40-3.97), being in the age groups 18-24 years (aOR 2.28, 95% CI: 1.03-5.01) or 25-29 years (aOR 3.06, 95% CI 1.35-6.93) compared to those aged 30 or older, having higher education levels, such as completing secondary education (aOR 1.76, 95% CI 1.01-3.06) or university education (aOR 2.65, 95% CI 1.18-5.96) in contrast to having no education. Lastly, perceiving a benefit from PrEP (aOR 9.52, 95% CI 4.27-21.22), and engaging in vaginal sex with a woman using a condom in the last 12 months (aOR 1.82, 95% CI 1.14-2.91) versus not. Impressively, 96% of participants expressed a strong willingness to use PrEP.

**Conclusion:** Among Rwandan MSM, there is a high level of awareness of PrEP, notably associated with factors such as residing outside Kigali, younger age, higher education, perceived benefits of PrEP and condom use during vaginal sex in the past year. Furthermore, a significant portion of participants demonstrated an intense desire to use PrEP, suggesting promising possibilities for its extensive implementation among this group of people. The findings from this study emphasize the importance of implementing highly focused awareness campaigns, personalized intervention, and comprehensive sexual health education programs in order to enhance the adoption of PrEP and bolster HIV prevention initiatives among the Rwandan population of MSM

## INTRODUCTION

Pre-exposure prophylaxis (PrEP) is highly effective in preventing new Human Immunodeficiency Virus (HIV) infections (1) and is recommended by the World Health Organization (WHO) as a key prevention strategy (2, 3). Sexual and gender minority (SGM) men who have sex with men (MSM) in sub-Saharan Africa (SSA) are disproportionately impacted by HIV, with HIV prevalence ranging from 6% to 17% (4,5,6). The prevalence of HIV among MSM in SSA is influenced by the legal and social environment, where in some countries homosexuality is considered a crime attracting severe penalties (7).

The Integrated Biological and Behavioral Surveillance Survey (IBBSS) conducted in 2020 revealed that the overall HIV prevalence among MSM in Rwanda was 4.3%, with the highest rate in Kigali City at 11.3% (5). Furthermore, a study conducted in the same year among MSM and transgender women in Kigali reported an HIV prevalence of 10% (8). This indicates that the prevalence of HIV infection among MSM in Rwanda is two to three times higher than in the general population (9,10,6) thus necessitating additional HIV preventive interventions in this vulnerable group. Rwanda has recognized PrEP as a crucial element of its HIV prevention strategy (11). However, despite a country-wide rollout of PrEP in 2019 (12), there is limited information on its awareness, willingness to use it, access to it, and its utilization among key populations like MSM.

Awareness of PrEP among MSM varies between countries due to different economic statuses and supporting policies (13). In SSA, PrEP awareness among MSM is generally relatively low, ranging from 20 to 60% (14, 15, 6, 16) compared to other key populations. PrEP awareness varies by demographic characteristics such as education, age, and migration status (17) whereas willingness to use PrEP varies by country and survey time and ranges between 32-92%, suggesting a variety of factors associated with the likelihood of using it (18). Despite this, there are encouraging signs of increasing awareness as more African countries implement PrEP projects (19). However, challenges such as limited availability (20), and access (21) persist, underscoring the need for data on factors determining awareness of and willingness to use PrEP among at-risk MSM (22).

A Kenyan study revealed that individual and interpersonal factors such as condom use self-efficacy, perceived ability to use PrEP, and membership in a Lesbian, Gay, Bisexual, Transgender (LGBT) organization, contributed significantly to the awareness of and willingness to use PrEP (16). We previously described awareness of and willingness to use PrEP in MSM in Rwanda, reporting that nearly half (48%) of MSM sampled for the study were aware of PrEP, and most (83%) were willing to use it (6). However, the sample was limited to a cohort of MSM living in the City of Kigali, therefore, it was not representative of MSM in the country as a whole, thus, a comprehensive survey was required to include MSM residing outside of Kigali. In addition, for this cohort study, participants were to be enrolled through snowball sampling and community-based MSM organizations, their HIV status remained unknown, and they were required to provide informed consent to undergo HIV testing (23,10,6). The objective of the present study using an online survey was to assess awareness of and willingness to use PrEP among a more representative population of Rwandan MSM. Highlighting these aspects can guide efforts to enhance awareness and accessibility to PrEP in this vulnerable population.

## METHODS

### Study design, setting, and population

We conducted a cross-sectional study in Rwanda using an anonymous online survey between April and June 2019. Ethics approval for the study was granted by the Rwanda National Ethics Committee and the Albert Einstein College of Medicine Institutional Review Board. We collaborated with leaders from MSM organizations in Kigali who participated in prior research (23, 6) to form a Community Advisory Board (CAB) for the study. MSM organizations focus on specific human rights issues for members of their community and, in collaboration with various partners, provide HIV and Sexually Transmitted Infection (STI) testing and prevention support and linkage to care for members of their community who need those services. The CAB facilitated the testing and validation of the study questionnaire, provided information about the study, and supported the distribution of the survey link to community members as needed.

Eligibility criteria for participants included: age ≥18 years, self-identify as MSM, residence in Rwanda, self-reported engagement in receptive or insertive anal sex in the last 12 months, and self-reported HIV-negative serostatus. We excluded participants from a prior and ongoing cohort study that we are undertaking because they were exposed to PrEP-related information during the study visits (23). The language of the survey questionnaire was Kinyarwanda, which is widely spoken in Rwanda.

### Data collection

The questionnaire was piloted with members of the CAB and subsequently revised before distribution to eligible participants. Using Qualtrics (24), web-links generated were sent through WhatsApp or by e-mail. It was requested that CAB members disseminate the survey web URLs to individuals within their social network and to members of their organizations. Upon obtaining the survey link, participants were requested to respond to the screening survey questions in order to ascertain their eligibility. If found to be eligible, they proceeded to complete a digitized informed consent form prior to commencing the survey (Figure 1). In addition, the survey was distributed by the participants who responded, as the last question asked them to share the survey link with their friends and social networks that they knew or felt were MSM. Participants who completed the survey were given an incentive of 2,500 Rwandan francs (∼$2), which they received through mobile money transfers to their cell phones.

**Figure 1.**
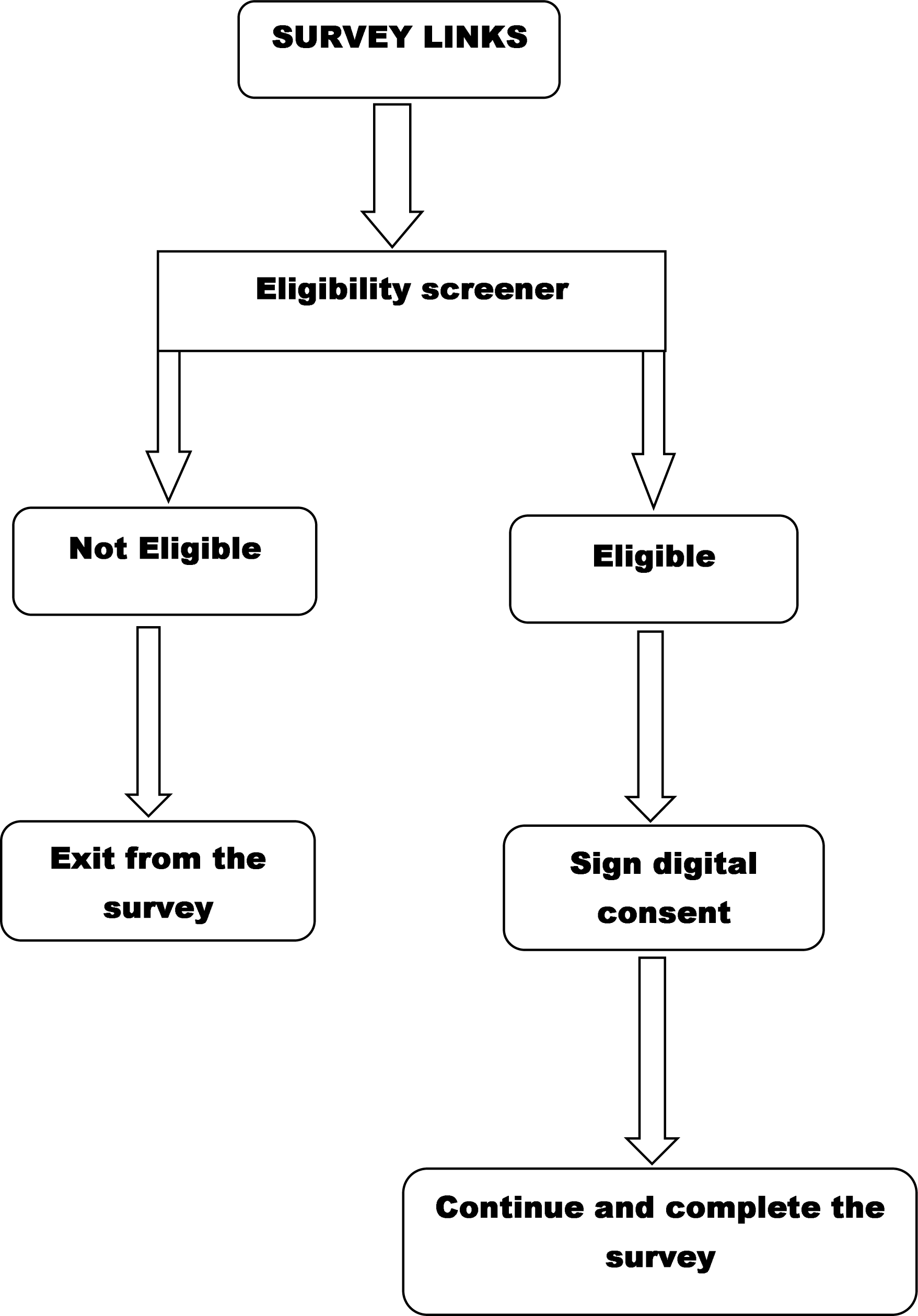
Process for participating in the study survey.

### Outcomes and Measures

We assessed two main outcomes, 1) awareness of PrEP, which was preceded by an introductory text as follows: “*PrEP is a new way to prevent getting infected with HIV. PrEP is a pill taken once a day by an HIV-negative person to protect themselves from getting HIV BEFORE they have sex. You can stop using the medicine at any time, in consultation with a doctor/nurse/healthcare worker. PrEP is extremely effective and safe*”. Following this introduction, participants were asked: “*Before today, how much would you say you knew about PrEP*?” Responses were categorized as follows: “*I have never heard of PrEP, I’ve heard about it, but I did not know what it is, I know a little bit about it, I know a fair amount about it, I know a lot about it*”. 2) Willingness to use PrEP, by asking: “*If PrEP were available for free, would you be willing to start taking PrEP in the next 1 month to protect yourself from getting HIV*”? Responses were categorized as “*yes, no, not sure*”.

Independent variables included: 1) Socio-demographic characteristics including age (18-24, 25-29, 30 years and above), gender at the time of the survey (man, transgender, intersex, female), place of residence (Kigali, another place in Rwanda), highest educational level completed (no education, primary, secondary, vocational, and university) and income (no income, < 150, 000 RWF, and ≥ 150, 000 RWF), 2) General and sexual health status including time since last consultation with a healthcare provider (never, less than six months, six months to one year, and more than one year ago), willingness to receive information about sexual health or to stay health through social media was included, categorized as no versus yes, 3) Sexual behaviors including current relationship status (not in a relationship versus being in a relationship with a man, a woman or a transgender), disclosure of sexual identity (to friends and/or colleagues, family members and healthcare providers), having anal sex without a condom with another man or woman in the last 12 months, 4) perceived benefit in taking PrEP by asking : “*Do you think you might benefit from PrEP*” with Yes versus No response options.

## Data analysis

Data were analyzed using IBM SPSS statistics (version 21). Frequencies and proportions were generated from categorical variables. Responses for variables on PrEP awareness were dichotomized as “**Yes**” (“I’ve heard about it, but I did not know what it is, I know a little bit about it, I know a fair amount about it, I know a lot about it”) and, “**No**”(“I have never heard about PrEP”). For willingness to use PrEP, responses were dichotomized as either **Yes** or **No (**“no, not sure”). Gender was analyzed as man vs. other gender, while the educational level was analyzed as a three-category variable (no education or completed primary, completed secondary or vocational training, completed university). A logistic regression analysis was used to determine the statistical significance of the relationship between independent variables and study outcomes in a bivariate analysis. For multivariable analysis, only independent variables with a p-value < 0.05 in the bivariate analysis were included. The adjusted odd ratios (aOR) and 95% confidence interval (CI) were used to present the associations between independent and outcome variables.

### Data flow

A total of 3,053 clicks were recorded for the survey, out of which 2,465 were excluded due to ineligibility. Additionally, 67 response clicks were excluded because respondents either completed the survey in an unusually short time (<2 minutes, 61 cases) or took an excessively long time (>300 minutes, 1 case) and because age information was not reported in 5 cases. Consequently, the final analytic sample comprised 521 participants (see Figure 2).

**Figure 2.**
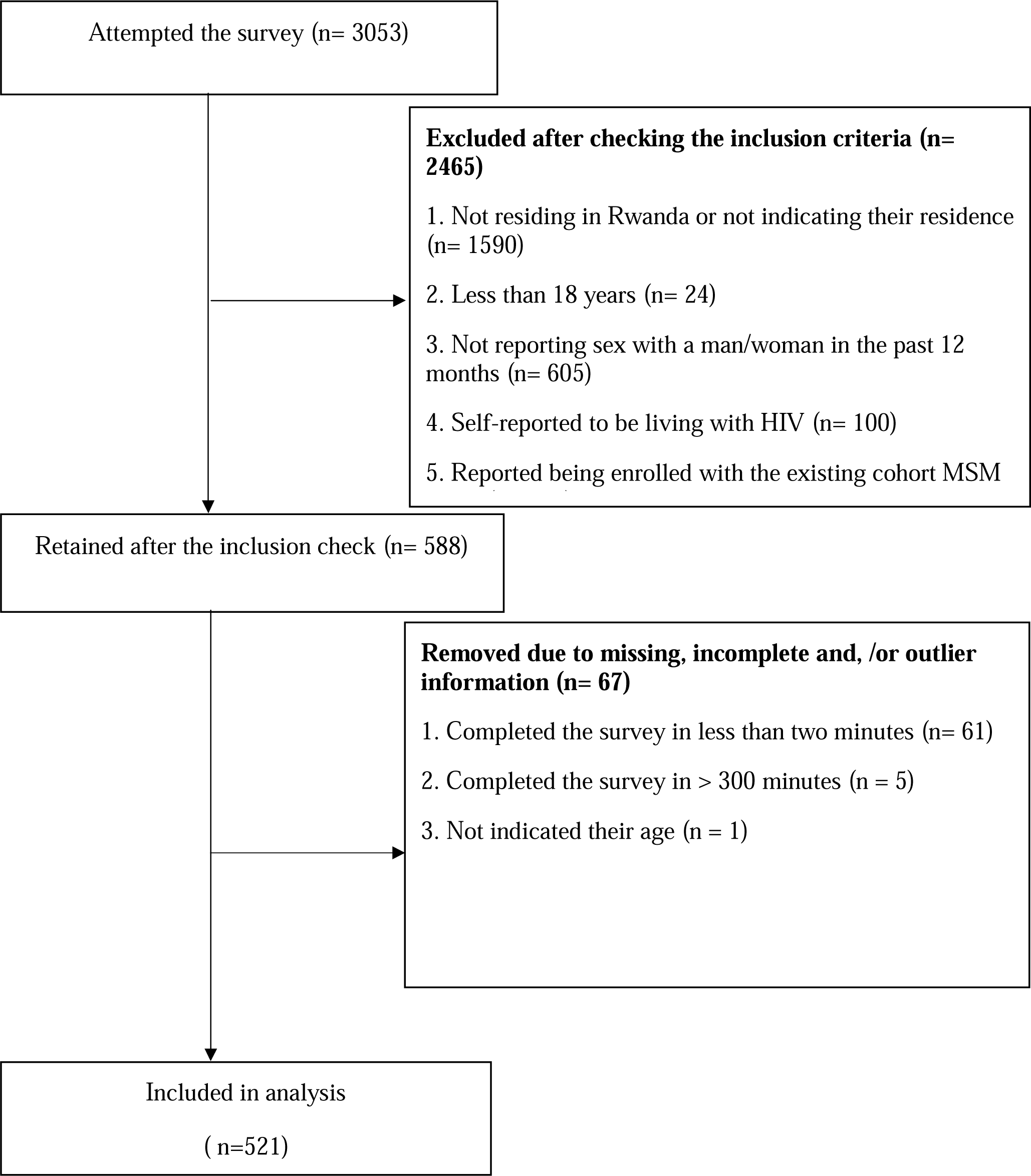
Data flow.

## RESULTS

### Socio-demographic characteristics of participants

Among the 521 participants included in this analysis, 318 (61%) reported living in the City of Kigali in the past 6 months, while 203 (39%) resided outside of the City of Kigali during the same period. Furthermore, 330 of 521 (63%) participants were 24 years of age or younger, and 374 (72%) self-identified as men at the time of the survey. Additionally, 319 (62%) reported having completed either secondary or vocational education, while 379 (73%) indicated they had no monthly income (Table 1).

**Tables 1.** Characteristics and Descriptions of Study Participants.

| **Variable** | **Frequency** | **Percent** |
| --- | --- | --- |
| **Living area in the past 6 months** |  |  |
| Kigali | 318 | 61 |
| Another place in Rwanda | 203 | 39 |
| **Age of respondents** |  |  |
| 18-24 years | 330 | 63.3 |
| 25-29 years | 141 | 27.1 |
| 30 years and above | 50 | 9.6 |
| **Current gender identity** |  |  |
| Male | 374 | 71.8 |
| Others | 147 | 28.2 |
| **Highest level of education completed** |  |  |
| None or primary | 120 | 23.4 |
| Secondary + vocational | 319 | 62.3 |
| University | 73 | 14.3 |
| **Monthly income** |  |  |
| No income | 379 | 73.3 |
| < 150000 RWF | 125 | 24.2 |
| ≥ 150000 RWF | 13 | 2.5 |
| **Most recent consultation with a healthcare provider** |  |  |
| Never | 23 | 4.4 |
| Less than 6 Months | 291 | 55.9 |
| 6 months to 1 year | 131 | 25.1 |
| More than 1 year ago | 76 | 14.6 |
| **Willingness to receive information for staying health on WhatsApp, Facebook Messenger, SMS, E-mail, or on another program on your mobile phone** |  |  |
| Yes | 483 | 94.7 |
| No | 27 | 5.3 |
| **You think you might benefit from PrEP** |  |  |
| Yes | 471 | 91.1 |
| No | 46 | 8.9 |
| **Current relationship status** |  |  |
| In a relationship ( man, female or transgender) | 101 | 19.6 |
| Not in relation with some one | 415 | 80.4 |
| **Disclosure of sexual attractions to friends and/or colleagues/family member or providers** |  |  |
| Yes | 74 | 35.1 |
| No one knows | 137 | 64.9 |
| **Having anal sex without a condom with another man in the last 12 months** |  |  |
| Yes | 234 | 47.6 |
| No | 258 | 52.4 |
| **Having sex intercourse without a condom with a woman in the last 12 months** |  |  |
| Yes | 218 | 43.9 |
| No | 279 | 56.1 |
| **Willingness to receive information about sexual health on WhatsApp, Facebook Messenger, SMS, E-mail, or on another program on your mobile phone** |  |  |
| Yes | 479 | 93.4 |
| No | 34 | 6.6 |
| **Awareness of PrEP** |  |  |
| I know about PrEP | 381 | 73.1 |
| I have never heard of PrEP | 140 | 26.9 |
| **Willingness to use PrEP** |  |  |
| Willing to use | 498 | 95.6 |
| Not willing to use | 23 | 4.4 |

### General, sexual health, behavior and acceptability of technology to receive health information

Over half (291/521) of the participants (56%) reported having had a health consultation with a healthcare provider in the six months prior to survey completion. Nearly all (483 or 95% and 479 or 93%) of the participants reported that they were willing to receive information about staying healthy or sexual health using Whatsapp, Facebook Messenger, SMS, e-mail or any other virtual method.

About 415/521 (80%) reported that they were not in any sexual relationship at the time of survey. One hundred and thirty-seven respondents (65%), reported that they have not disclosed their sexual orientation to anyone. Condom use in the past 12 months was reported by 258 (52%) of those who had anal sex with other men and 279 (56%) of those who had vaginal sex. In addition, most participants 471 (91%) thought that PrEP could benefit them.

### Awareness of and willingness to use PrEP

Overall, 381 (73%) participants reported awareness of PrEP at the time of the survey. In the bivariate analysis, awareness of PrEP was found to be significantly associated with various social demographic, general, sexual health, and behavior variables among participants (Table 2). In the multivariable logistic regression analysis, individuals who reported living elsewhere in Rwanda were almost twice as likely to be aware of PrEP (aOR 2.36, 95% CI 1.40-3.96) compared with those living in the City of Kigali. PrEP awareness was significantly higher among participants aged 18-24 years (aOR 2.28, 95% CI 1.03-5.01) and 25-29 years (aOR 3.06, 95% CI 1.36-6.93) compared to those aged ≥30 years. Similarly, participants with secondary/vocational education (aOR 1.77, 95% CI 1.01-3.07) and university education (aOR 2.65, 95% CI 1.18-5.97) reported higher awareness of PrEP compared to those with no education or only primary education. Additionally, awareness of PrEP was significantly higher among participants who perceived a benefit from PrEP (aOR 9.52, 95% CI 4.28-21.22) compared to those who did not. Furthermore, those reporting vaginal sex with a condom were more aware of PrEP (aOR 1.82, 95% CI 1.14-2.91) than those reporting vaginal sex without a condom. This is described in Table 3. Overall, 498 (96%) of participants were willing to start taking PrEP in the next one month to protect themselves from getting HIV (Table 1).

**Table 2.**
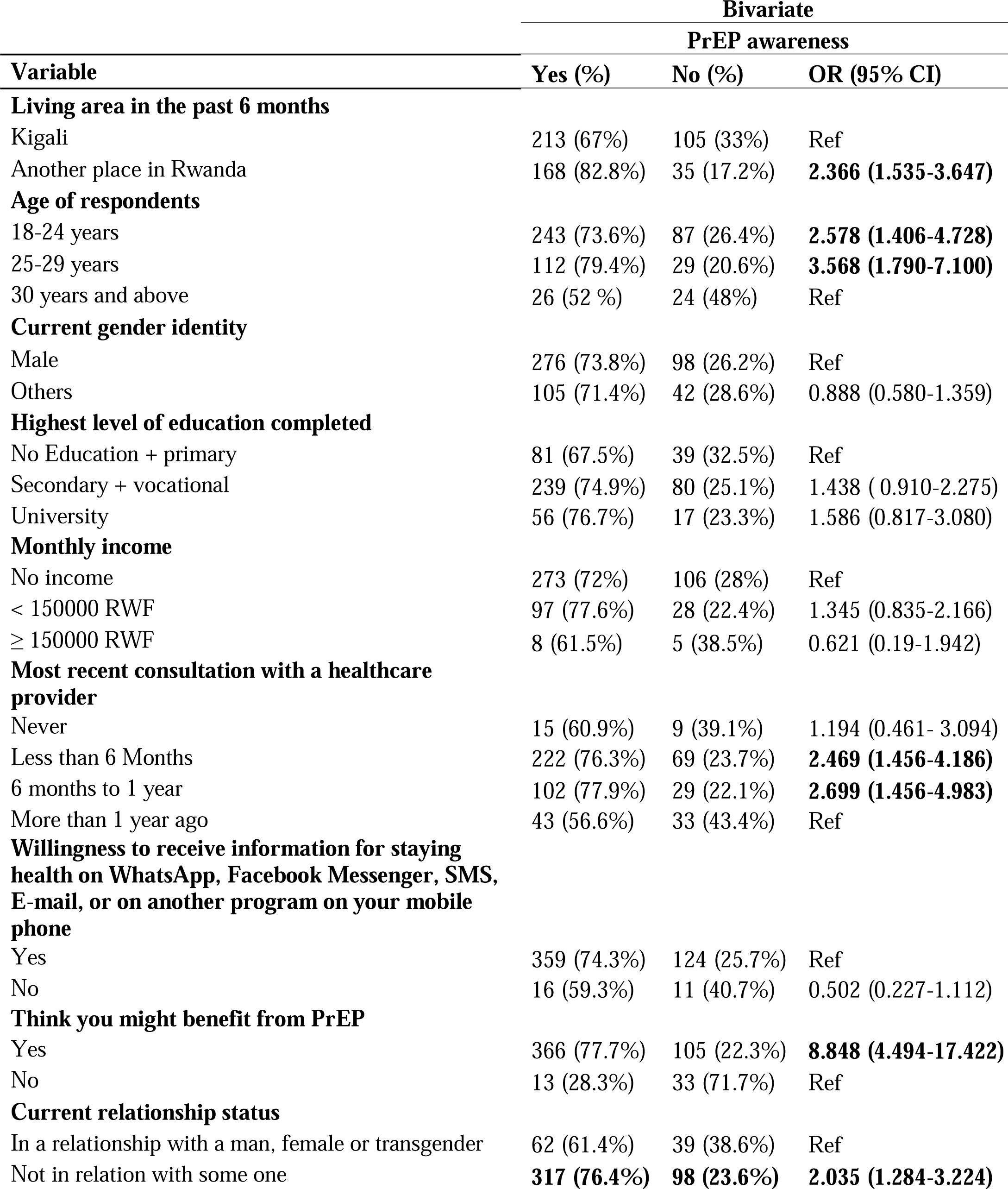
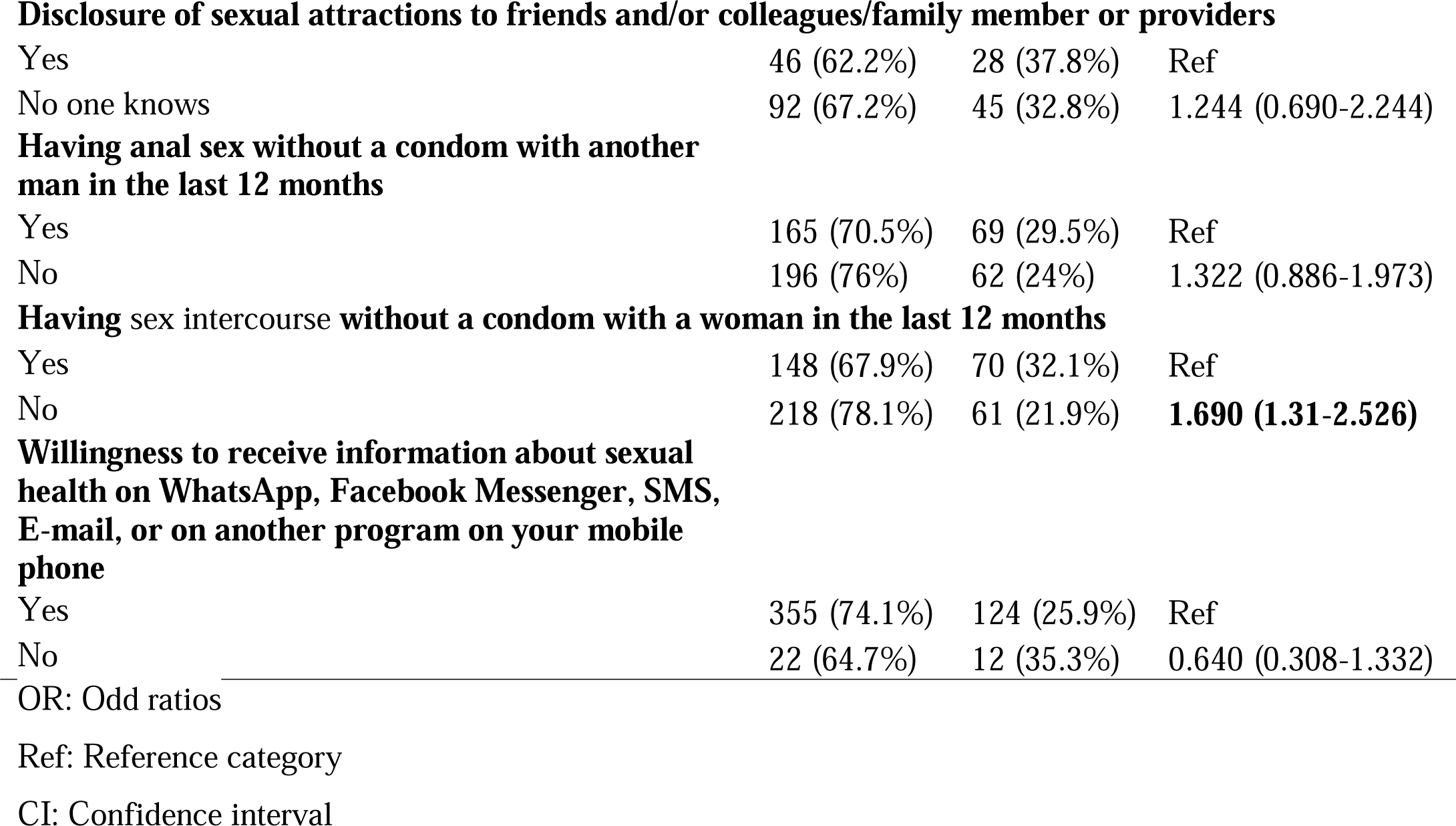
Bivariate analysis of PrEP awareness among Rwandan MSM.

**Table 3.** Multivariable analysis of PrEP awareness among Rwandan MSM.

|  | Multivariable |  |  |  |
| --- | --- | --- | --- | --- |
|  | PrEP awareness |  |  |  |
| Variable | aOR | 95% C.I. |  | P-value |
|  |  | Lower | Upper |  |
| Living area in the past 6 months |  |  |  |  |
| Kigali |  |  |  |  |
| Another place in Rwanda | 2.359 | 1.404 | 3.963 | 0.001 |
| Age of respondents |  |  |  |  |
| 18-24 years | 2.281 | 1.038 | 5.014 | 0.04 |
| 25-29 years | 3.067 | 1.358 | 6.926 | 0.007 |
| 30 years and above | Ref |  |  |  |
| Highest level of education completed |  |  |  |  |
| None or primary | Ref |  |  |  |
| Secondary + vocational | 1.764 | 1.015 | 3.067 | 0.044 |
| University | 2.658 | 1.184 | 5.966 | 0.018 |
| Most recent consultation with a healthcare provider |  |  |  |  |
| Never | 0.872 | 0.29 | 2.626 | 0.808 |
| Less than 6 Months | 1.605 | 0.842 | 3.06 | 0.151 |
| 6 months to 1 year | 1.763 | 0.853 | 3.644 | 0.126 |
| More than 1 year ago | Ref |  |  |  |
| Think you might benefit from PrEP |  |  |  |  |
| Yes | 9.529 | 4.279 | 21.22 | < 0.0001 |
| No | Ref |  |  |  |
| Current relationship status |  |  |  |  |
| In a relationship with a man, female or transgender | Ref |  |  |  |
| Not in relation with some one | 1.747 | 0.96 | 3.177 | 0.068 |
| Having sex intercourse without a condom with a woman in the last 12 months |  |  |  |  |
| Yes | Ref |  |  |  |
| No | 1.826 | 1.145 | 2.91 | 0.011 |

## DISCUSSION

The anonymous online survey conducted among MSM in Rwanda has revealed significant findings related to the awareness and willingness to use PrEP as an HIV infection prevention method. The study sample demonstrated a substantial level of awareness of PrEP, with the majority of participants expressing a strong willingness to initiate PrEP within the next month as a protective measure against HIV infection. Furthermore, a significant proportion of the participants mentioned having had recent medical consultations and exhibited a strong willingness towards obtaining health-related information via diverse online platforms.

While our study reported high PrEP awareness, it is important to note that awareness levels have varied in previous research conducted in low- and middle-income countries (LMICs) (25,26, 27, 28, 29, 30, 31), including our previous study limited to MSM living in the City of Kigali (6). This variability can be attributed to factors such as the clarity of PrEP definition provided to participants, the timing of the survey compared to PrEP availability (12, 32), and the broader trends in PrEP awareness over time (25). Rwanda’s inclusion of PrEP in national HIV prevention guidelines in 2018 (32, 12) may explain the higher awareness in our study compared to previous publications.

A high level of PrEP awareness is an indication that public health campaigns aimed at HIV/AIDS prevention are working. Knowledgeable people are more likely to actively seek information, speak with medical professionals, and make informed choices, which could lead to an increase in the use of PrEP among populations that are at risk (33,34,35). This awareness also enables public health organizations to target educational campaigns effectively, directing resources to populations with lower awareness to promote equitable access to PrEP information and hence potential use.

Our results indicate that awareness of PrEP was higher among MSM who reported residing outside Kigali in the past 6 months than among those who reported living in Kigali City. This suggests that factors beyond awareness campaigns and information access, such as individual living independence, may influence PrEP awareness. Mobility among MSM across Rwanda, as observed in a prior report (36), may contribute to this difference. A study conducted in high-income settings, including Atlanta, Chicago, and New York City, similarly documented disparities in PrEP awareness associated with place of residence and geographic location (37). Contrary to our findings, a study conducted on gay, bisexual, and other men who have sex with men (GBMSM) in Nigeria, suggested that those living in urban areas may exhibit higher levels of awareness (38), our results indicated a different trend. Further research and in-depth data analysis are imperative to comprehensively discern the underlying factors contributing to the disparity in PrEP awareness among MSM in various regions of Rwanda.

Additionally, our research revealed that high levels of awareness were associated with younger ages and higher educational attainment, which is in accordance with Rwanda’s demographic trends (5, 6), and similar findings in SSA (39,40, 41, 42, 25). These results highlight the need for targeted outreach and education efforts tailored to different age groups, particularly younger individuals who may be more receptive to new prevention methods. In addition, our findings also suggest that public health education and awareness campaigns should consider the educational background of the target audience to effectively disseminate information about PrEP.

Furthermore, participants who perceived a benefit from PrEP demonstrated substantially higher awareness of it, highlighting the importance of emphasizing PrEP’s advantages and effectiveness in HIV prevention in public health campaigns. The perception of PrEP benefits has consistently been identified as a factor facilitating its potential use among MSM (28).

An interesting finding was the association between condom use during vaginal sex and PrEP awareness. Those reporting condom use were more aware of PrEP, suggesting a connection between safe sex practices and knowledge about PrEP. Individuals who use condoms may consistently already have higher awareness of HIV prevention methods, making them more receptive to information about PrEP. Also, healthcare providers or sexual health education programs may be more likely to discuss PrEP with individuals who prioritize safe sex practices, leading to increased awareness. This finding highlights the significance of comprehensive sexual health education and outreach initiatives that provide information on PrEP in addition to contraceptive use or other protective measures promotion. Similarly, in a neighboring country, Kenya, higher condom use and self-efficacy have been reported to be associated with PrEP awareness (15).

Finally, the study revealed a high level of willingness among respondents to use PrEP, indicating readiness for PrEP implementation among MSM in Rwanda, in line with broader trends observed among MSM in sub-Saharan Africa (15,28, 43, 44).

### Limitations

The survey’s anonymity and the use of snowball sampling to gather respondents may have contaminated responses from some participants who lived in the same community as individuals who had previously participated in another cohort study testing PrEP awareness (6). Furthermore, the limited number of online-based studies conducted with MSM in SSA restricts the effective comparison of these results across the region.

Finally, the overwhelming willingness of nearly all respondents to use PrEP prevented us from conducting a bivariate or multivariate analysis to identify potential associations between participants’ socio-demographic, health, and/or sexual health characteristics and their observed willingness to use PrEP.

### Conclusion

The results of the anonymous online survey indicate a high level of awareness of PrEP among MSM, with around two-thirds being aware of PrEP. Significantly, the vast majority of participants indicated a readiness to commence PrEP use within the following month as a preventive measure against HIV. A wide range of socio-demographic variables were found to be correlated with PrEP awareness, such as place of residence, age, level of education, perceived benefit from PrEP, and condom use. This study provides crucial information regarding Rwandan MSM’s awareness of PrEP and propensity to use it. This underscores the importance of implementing focused awareness campaigns, individualized interventions, and comprehensive sexual health education in order to encourage the use of PrEP and prevent HIV infection among this demographic. Policy-makers should design and implement targeted PrEP awareness campaigns that take into account the demographic variations in awareness identified in the survey. Special emphasis should be placed on reaching older MSM individuals and those with lower levels of education to ensure equitable access to information about PrEP. Highlighting the advantages of PrEP in HIV prevention can encourage more individuals to consider and use it as a preventive measure.

## Data Availability

All data produced in the present study are available upon reasonable request to the authors

## Author Contributions

**KA, VP, GM, AA** contributed substantially to the development of the study protocol**, AM** implemented the study and led the drafting of the manuscript, **AM, VP, GM, EN, LN KA, JR, and AA** contributed substantially to the study design and editing of the manuscript for important intellectual content. **AM, QS, NG, JR, VP** did the study analysis and data interpretation for the work, and **GK, AP, JR** contributed substantially to the drafting of the work and its revision. All authors have contributed to the writing of the manuscript and have approved the submitted version.

## Acknowledgment

We thank the Community Advisory Board Members who provided support during the pilot testing of the study survey, research participants, and Research for Development (RD-Rwanda) for their support.

